# County-level exposures to greenness and associations with COVID-19 incidence and mortality in the United States

**DOI:** 10.1101/2020.08.26.20181644

**Authors:** Jochem O Klompmaker, Jaime E Hart, Isabel Holland, M Benjamin Sabath, Xiao Wu, Francine Laden, Francesca Dominici, Peter James

## Abstract

**Background:** COVID-19 is an infectious disease that has killed more than 246,000 people in the US. During a time of social distancing measures and increasing social isolation, green spaces may be a crucial factor to maintain a physically and socially active lifestyle while not increasing risk of infection.

**Objectives:** We evaluated whether greenness is related to COVID-19 incidence and mortality in the United States.

**Methods:** We downloaded data on COVID-19 cases and deaths for each US county up through June 7, 2020, from Johns Hopkins University, Center for Systems Science and Engineering Coronavirus Resource Center. We used April-May 2020 Normalized Difference Vegetation Index (NDVI) data, to represent the greenness exposure during the initial COVID-19 outbreak in the US. We fitted negative binomial mixed models to evaluate associations of NDVI with COVID-19 incidence and mortality, adjusting for potential confounders such as county-level demographics, epidemic stage, and other environmental factors. We evaluated whether the associations were modified by population density, proportion of Black residents, median home value, and issuance of stay-at-home order.

**Results:** An increase of 0.1 in NDVI was associated with a 6% (95% Confidence Interval: 3%, 10%) decrease in COVID-19 incidence rate after adjustment for potential confounders. Associations with COVID-19 incidence were stronger in counties with high population density and in counties with stay-at-home orders. Greenness was not associated with COVID-19 mortality in all counties; however, it was protective in counties with higher population density.

**Discussion:** Exposures to NDVI had beneficial impacts on county-level incidence of COVID-19 in the US and may have reduced county-level COVID-19 mortality rates, especially in densely populated counties.

## Introduction

The global spread of Severe Acute Respiratory Syndrome Coronavirus 2 (SARS-CoV-2), the virus responsible for COVID-19, has caused a worldwide public health emergency. (Sohrabi et al. 2020; WHO 2020a) The outbreak was declared a pandemic by the World Health Organization (WHO) on March 11, 2020, (WHO 2020b) and as of November 16, 2020, 54.5 million cases of COVID-19 had been documented worldwide, and more than 1.3 million deaths had been recorded. (Johns Hopkins Coronavirus Resource Center 2020) To date, there are few effective therapies and no vaccine, therefore public health measures at the population level (e.g. social distancing measures, stay-at-home orders, public education initiatives (Prem et al. 2020; Tammes 2020)) have been the primary approach for reducing transmission.

The coronavirus pandemic presents an unprecedented situation for the globe; however, this is not the first time the world has confronted a large-scale infectious disease threat. Historical approaches to combat infectious disease outbreaks provide crucial lessons that we can still apply today. One of those approaches is the use of urban parks as a resilience measure. Frederick Law Olmsted, who designed New York’s Central Park, Boston’s Emerald Necklace, and many other major urban parks, championed the concept of “parks as lungs” and he espoused “two great natural agents of disinfection: sunshine, and fall foliage”.(Beveridge and Hoffman 1997)

The spread of infectious diseases, like COVID-19, is dependent on the duration of infectiousness, transmissibility, and the contact rate.(Heederik et al. 2020; Delamater et al. 2019) These three factors generally summarize the basic reproduction number (R0). R0 is affected by numerous biological, socio-behavioral and environmental factors that influence pathogen transmission.(Delamater et al. 2019) Green spaces may influence the contact rate and in turn the reproduction number, as they provide a setting to obtain much needed physical activity and a place for social interactions while remaining the recommended safe distance (six feet). Because these activities take place outdoors, wind dilutes the amount of virus in the air substantially (Qian et al. 2020), which greatly decreases transmission risk. In addition, theory (Ulrich 1984; Kaplan & Kaplan 1989) and empirical evidence (Banay et al. 2019; Bezold et al. 2018) suggests that living near green spaces allows us to restore our attention and decrease stress, leading to lower incidence of depression, anxiety, and other negative psychological factors. During a time of social distancing measures and increasing social isolation, urban green spaces may be a crucial factor to maintain a physically and socially active lifestyle while not increasing risk of infection.

To quantify whether greenness is related to COVID-19 incidence and mortality, we compiled county level data on both greenness and COVID-19 outcomes. Furthermore, based on evidence that there are large disparities in incidence and mortality rates, we evaluated whether the relationship between greenness and incidence/mortality differed according to county-level population density, percentage of black residents, median home value and issuance of stay-at-home orders.

## Methods

Data used in this study are publicly available and links to each of the data sources can be found in Table S1.

### COVID-19 incidence and mortality data

The Johns Hopkins University Center for Systems Science and Engineering Coronavirus Resource Center provides daily updates about COVID-19 death counts and cases for each country.(Dong et al. 2020) For the US, county level data is provided by the US Centers for Disease Control and Prevention (CDC) and State governments. The number of COVID-19 cases is the sum of the number of deaths and active cases. As of April 14, 2020, CDC case counts and death counts included both confirmed and probable cases and deaths in accordance with CDC guidelines.(CDC 2020)

We downloaded data on the cumulative number of COVID-19 cases and deaths for each county through June 7, 2020. County-level COVID-19 mortality/incidence rates were defined as the ratio of COVID-19 deaths/cases to county level population size.(Wu et al. 2020)

### Greenness exposure

For each county, the Normalized Difference Vegetation Index (NDVI) was estimated using satellite imagery. The NDVI is calculated as the ratio between the red and near infrared values, and ranges from −1 to 1.(NASA 2020) Values close to 1 correspond to areas with complete coverage by live green vegetation, values close to zero correspond to areas without much live vegetation (e.g. rocks, sand) and negative values correspond to water. We used Landsat 8 (Collection 1 Tier 1 Operational Land Imager DN values, representing scaled, calibrated at sensor radiance (USGS 2020)) images for the entire US from April 1, 2020 up to May 31, 2020, to represent the exposure during the initial COVID-19 outbreak in the US. Landsat 8 images are generated every 16 days at 30m resolution. Using Google Earth Engine (https://earthengine.google.com/), cloud-free Landsat composites were created for the US. We calculated the spatially weighted mean April-May NDVI for each county in the US, after setting negative NDVI values to zero. For sensitivity analyses, we also used Landsat 8 images from June 1, 2019 up to August 31, 2019, to calculate the spatially weighted mean summer NDVI for each county. County shapefiles were based on the US Census Bureau Tiger dataset of 2018.(US Census Bureau 2020)

### Potential confounders

To adjust for potential confounding bias, we obtained data on several variables that might be linked to green space and COVID-19 incidence and mortality. We collected eleven county level Census variables from the 2000 Census (https://www.census.gov) and the 2010 5-year American Community Surveys (https://www.census.gov/programs-surveys/acs/): proportion of residents older than 65, proportion of residents aged 15-44, proportion of residents aged 45-64, proportion of Hispanic residents, proportion of Black residents, median household income, median home value, proportion of residents in poverty, proportion of residents with a high school diploma, population density, and proportion of residents that own their house. From the Behavioral Risk Factor Surveillance System (BRFSS) (https://www.countyhealthrankings.org/) we obtained the proportion of individuals that were obese and the proportion of current smokers in 2011, the most recent year available.

We used days since first COVID-19 case reported in a county as a proxy for stage of the COVID-19 outbreak. Further, we linked days since issuance of stay-at-home order (state-level), days since closure of non-essential businesses (state-level), and days since nursing home visitor ban (state-level) from the COVID-19 US State Policy Database (Raifman et al. 2020) to our data. Since the availability of adequate hospital resources might influence COVID-19 outcomes, we collected county-level information on the number of hospital beds available in 2019 from the Homeland Infrastructure Foundation-Level Data (HIFLD). In addition, we used state level information on number of COVID-19 tests performed up to June 7, 2020 from the COVID tracking project (https://covidtracking.com/).

Based on previous studies implicating relations between exposure to particulate matter less than 2.5 microns (PM_2.5_), temperature and/or relative humidity and COVID-19 incidence and mortality (Wu et al. 2020; Raines et al. 2020), we also adjusted our analyses for these factors. Temperature and relative humidity data were available from Gridmet, and we created long-term (2000-2016) summer (June-August) and winter (December-February) averages for each county.(Abatzoglou 2013) PM_2.5_ concentration estimates for 2000-2016 were derived from an established exposure prediction model.(Van Donkelaar et al. 2019)

### Statistical methods

We used negative binomial mixed models to evaluate associations of NDVI with COVID-19 incidence and mortality. We report mortality rate ratios (MRR) and incidence rate ratios (IRR), i.e., exponentiated effect estimates from the negative binomial mixed model, and 95% CI per 0.1 unit NDVI increase. To evaluate effects of potential confounders, we specified a series of models with increasing covariate adjustment. In model 1 we only included a population size offset and a random intercept by state. In model 2 we additionally adjusted for degree of urbanization. In model 3 we added all county-level SES covariates and BRFSS covariates. In model 4 we added date since first COVID-19 case reported, date since issuance of stay-at-home order for each state, number of hospital beds per unit population. In model 5 we additionally included temperature, relative humidity and PM_2.5_ (main model for COVID-19 mortality). The number of tests per unit population was added to model 6 (main model for COVID-19 incidence). We used a general additive mixed model with penalized cubic regression splines (with 2 degrees of freedom as the upper limit) to evaluate whether the association of NDVI with COVID-19 mortality and incidence was linear in the full cohort, in rural counties, and in urban counties. We carried out all analyses in R statistical software and performed model fitting using the lme4 package or the gamm4 package (for spline analyses).(Package “lme4”; Package “gamm4”)

We evaluated whether associations of NDVI with COVID-19 deaths and cases were modified by population density, proportion of black residents, median home value, and issuance of stay-at-home order by adding an interaction term to the model. Significance of interaction terms were tested by Chi-square tests between the models with and without the interaction terms. We hypothesized that associations of NDVI with COVID-19 incidence and mortality were stronger in densely populated counties, in counties with issuance of stay-at-home orders, in counties with higher proportions of black residents, and in counties with lower median home values.

We conducted several sensitivity analyses to assess the robustness of the associations. We evaluated associations of summer NDVI in the full population, and in urban and rural counties. We excluded 27 counties comprising the New York metropolitan area (n = 3,062), as this area experienced the most severe COVID-19 outbreak. We also conducted analysis excluding counties with 10 or fewer confirmed COVID-19 cases. We additionally added days since closure of non-essential businesses and days since nursing home visitor ban to our models. To evaluate the impact of potential spatial residual confounding, we additionally added longitude and latitude of the centroid of each county to the models. In addition, we used county averages of NDVI with negative values excluded (instead of set to zero).

## Results

Our study cohort consisted of 3,089 counties of which 2,297 counties reported more than 10 cases. The highest COVID-19 death rates were in New York, Illinois, Michigan, Florida, Louisiana, and California (Figure 1). COVID-19 incidence rates were more equally spread over the US. NDVI values were high along the West coast and in the South. The median COVID-19 death rate per 100,000 individuals was 2.8 and the median COVID-19 incidence rate per 100,000 individuals was 163.5 (Table 2). NDVI was moderately positively correlated with % Black, % smoke, and PM_2.5_, and weakly negatively correlated with median household income (Figure S1).

**Figure 1.**
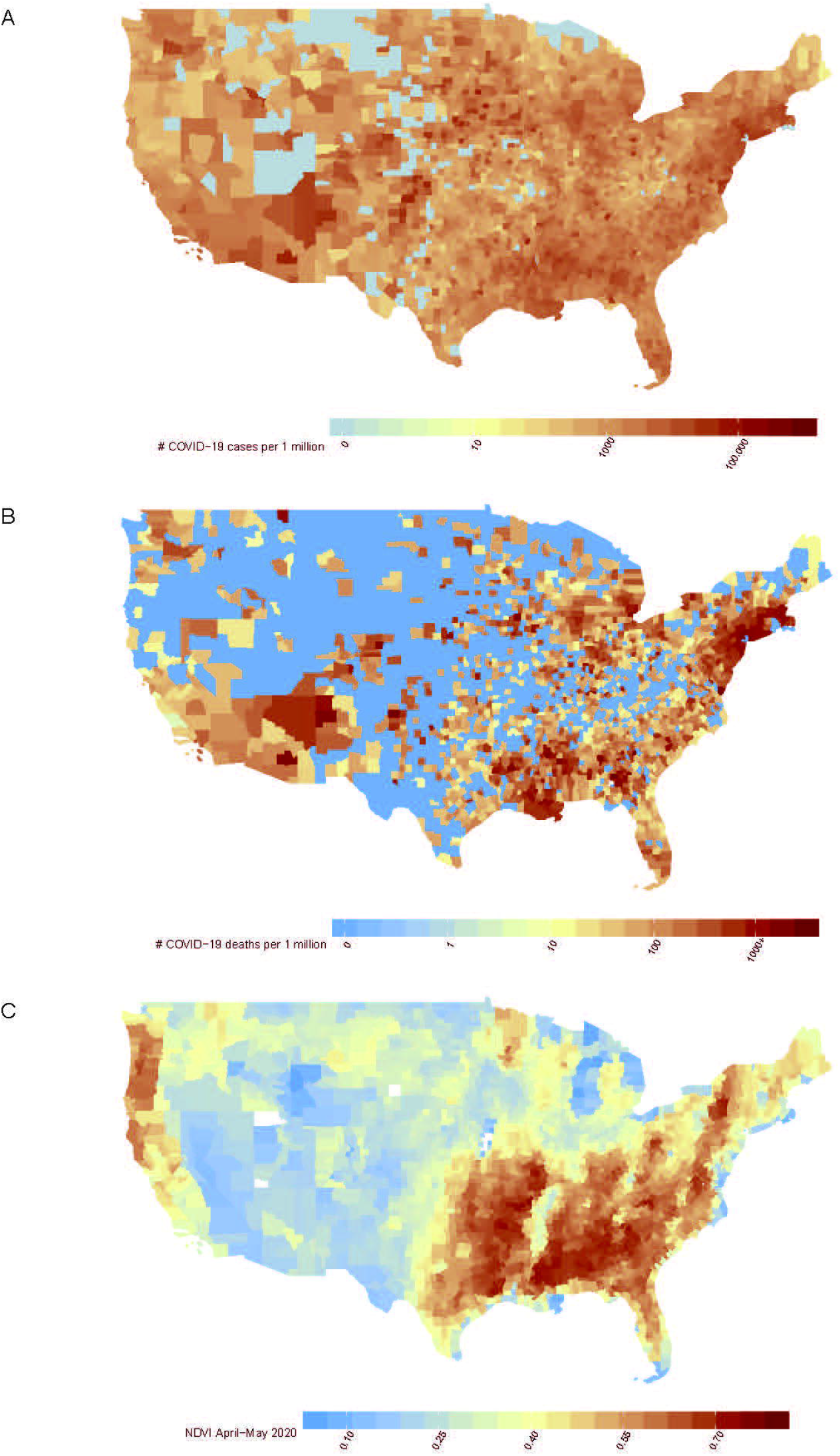
Maps of the US that show (A) the county-level number of COVID-19 cases per 1 million population in the United States up to and including June 7, 2020, (B) the county-level number of COVID-19 deaths per 1 million population in the United States up to and including June 7, 2020, and (C) county-level average NDVI (April-May 2020).

In main models (model 5 for mortality and model 6 for incidence) we found an IRR of 0.94 (95% CI: 0.90, 0.97) and a MRR of 0.99 (95% CI: 0.94, 1.05) per 0.1 unit increase in NDVI. There was little impact of population density on estimates; however, effects were attenuated in models that included county-level SES, BMI and smoking (Figure S2). The additional inclusion of epidemic stage, timing of stay-at-home-orders, hospital beds per capita, long-term exposures to PM_2.5_, weather and COVID test rate did not confound the association. Estimated IRR and MRR for all covariates included in the fully adjusted models can be found in Table S2. The overall exposure-response curve for COVID-19 incidence showed some small evidence of deviations from linearity, with a potential threshold effect around an NDVI of 0.5 (Figure 2). For urban counties, the curve for COVID-19 incidence was inverse and linear, while for rural counties increasing NDVI appeared beneficial at the lower end of the distribution only. Similar patterns, although slightly less pronounced, were observed for COVID-19 mortality.

**Figure 2.**
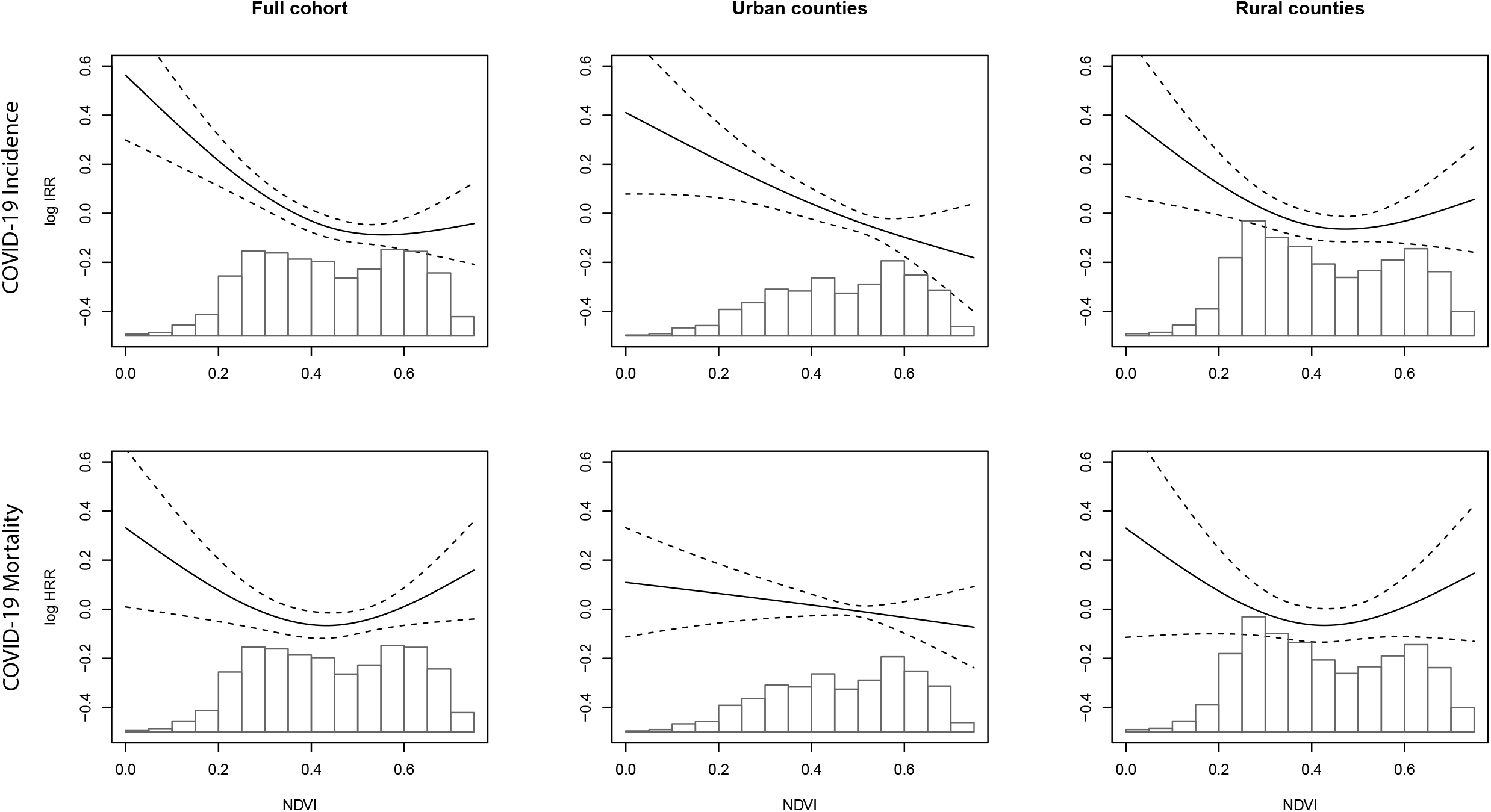
Exposure-response curves of the association of NDVI with COVID-19 incidence and COVID-19 mortality in the full cohort, in urban counties and in rural counties.

Associations of NDVI with COVID-19 incidence and mortality were positive in the lowest densely populated counties and negative in the highest densely populated counties (Figure 3). For COVID-19 incidence, but not for mortality, we found stronger associations for counties with higher median home values and issuance of stay-at-home orders. Associations of NDVI with COVID-19 incidence were similar across quintiles of the proportion Black residents, while we found a positive association with COVID-19 mortality in the lowest quintile.

**Figure 3.**
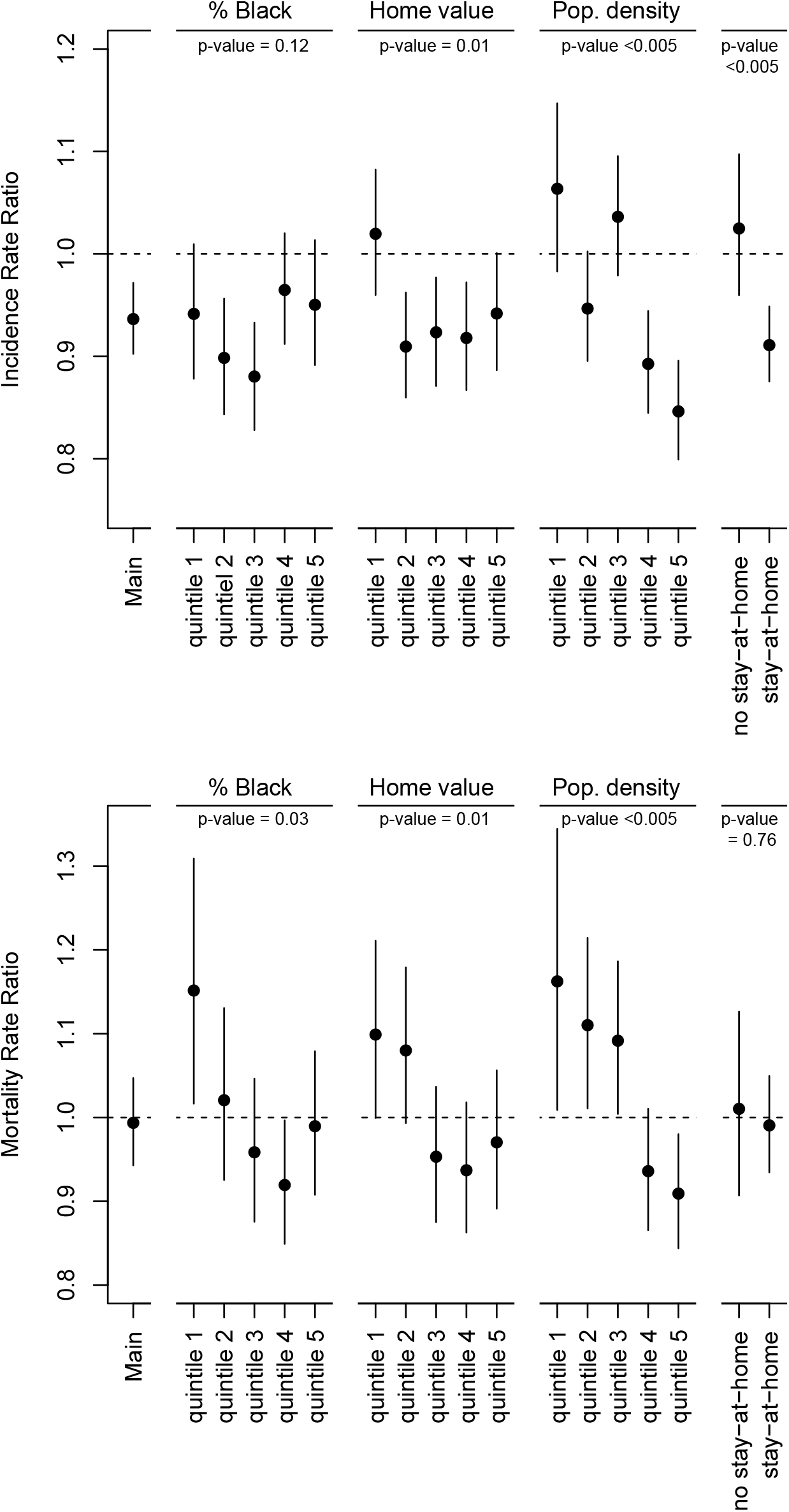
Associations of NDVI with COVID-19 incidence and COVID-19 mortality by strata. Main = main model, quintile 1 = lowest quintile, quintile 5 = highest quintile. Home value = median home value, Pop. density = population density, no stay-at-home = counties with no issuance of stay-at-home order, stay-at-home = counties with issuance of stay-at-home order ^a^. ^a^ Associations are expressed per 0.1 unit increase in NDVI.

In sensitivity analyses, associations were robust to additional adjustment for potential spatial clustering, days since closures of non-essential businesses or days since a nursing home visitor ban, exclusions of the NYC metro area, restriction to counties with at least 10 cases, or alternative procedure for calculating NDVI. (Figure S3). In the full cohort, associations of summer NDVI with COVID-19 incidence and mortality were weakly negative, but not-significant (Table S3). For urban counties, we found an IRR of 0.96 (95% CI: 0.91, 1.01) and a MRR of 0.94 (95% CI: 0.88, 1.00) per 0.1 unit increase in summer NDVI.

## Discussion

We observed that greenness in April-May of 2020 was inversely associated with COVID-19 incidence, especially in urban counties. An increase of 0.1 in NDVI was associated with a 6% decrease in COVID-19 incidence rate after adjustment for potential confounders at the county level. Associations with COVID-19 incidence were stronger in more densely populated counties, and in counties with stay-at-home orders. NDVI was not associated with COVID-19 mortality in all counties; however, NDVI was protective in counties with higher population density.

Several studies indicated that environmental exposures, such as air pollution, temperature, and humidity, could affect the spread and impact of infectious diseases because of their impact on host susceptibility and virus stability/survival. (Moriyama et al. 2020; Ciencewicki and Jaspers 2007; Martelletti and Martelletti 2020; Dowell and Shang Ho 2004) To the best of our knowledge, there are no studies that evaluated the effect of greenness on the spread and impact of infectious diseases. Since the COVID-19 outbreak, social distancing policies and guidelines have led to more time spent at home and therefore people may be more dependent on their immediate surroundings. Because gyms were closed in large parts of the US during this time period, people may have relied on parks to be physically active. Parks also provide places for social gatherings outdoors while remaining the recommended safe distance (six feet). Being outside might substantially reduce the chance of SARS-CoV-2 transmission. According to a study performed among 7,324 identified cases in China, only a single small outdoor outbreak was identified.(Qian et al. 2020).

We found stronger associations between NDVI and COVID-19 incidence. This seems plausible as neighborhood green space might affect contact rates and therefore COVID-19 incidence, while COVID-19 mortality also depends on available treatments and on host susceptibility, such as age and presence of chronic diseases. Several reviews showed inverse associations of greenness with a variety of diseases.(Fong et al. 2018; James et al. 2015) A couple of studies also reported inverse associations with cardiovascular and respiratory disease mortality, even after adjustment for air pollution.(Crouse et al. 2017; Vienneau et al. 2017) This suggests that increased amounts of greenness could influence host susceptibility. For COVID-19 incidence, associations were stronger (and linear) in urban versus rural counties. This is in line with the literature on the health effects of green spaces, which suggest benefits of green space are stronger in urban areas.(Fong et al. 2018) In urban areas, vegetation likely represents urban parks and street greenery, which are generally accessible and suitable spaces for recreational activities. This may not be true for vegetation in rural areas.

Associations of April-May NDVI differed a bit from associations of summer NDVI. Summer NDVI was weakly, but not significantly, associated with COVID-19 incidence and mortality. April-May NDVI was stronger associated with COVID-19 incidence, while summer NDVI was slightly stronger associated with mortality. We speculate that summer NDVI might better capture the long-term impact of greenness on health and therefore the impact of greenness on host susceptibility, while April-May NDVI might better capture the impact of greenness on contact rates as it largely overlaps with the beginning of the COVID-19 outbreak.

For COVID-19 incidence, we found stronger associations in densely populated counties and counties with high median home values. Median home value is likely related to health insurance and the ability to work from home, which affects COVID-19 incidence. The positive associations of NDVI with COVID-19 mortality in the lowest population density quintiles could be because an increase in greenness in these areas is related to limited access to health care. Associations of NDVI with COVID-19 incidence were modified by state-level issuance of stay-at-home orders.

Individuals living in states with stay-at-home orders might spend more time at home and are thus more dependent on their immediate surroundings, like greenness. Individuals living in states without stay-at-home orders might not practice social distancing and may differ in COVID-19 health risk perceptions. However, differences in associations could also be due to differences in epidemic stage (number of COVID-19 cases) in counties with and without stay-at-home orders. Associations of NDVI with COVID-19 mortality, but not COVID-19 incidence, were modified by percentage Black. NDVI was harmful in the counties with the lowest proportion of Black residents, but not in other quintiles. This finding may be related to higher observed rates of mortality among Black individuals, or may reflect the moderate correlation between percentage of Black residents and population density.

This study has several strengths. We used NDVI for April-May 2020, largely overlapping with the beginning of the COVID-19 outbreak in the US, allowing us to assess the impact of temporally relevant exposures on incidence and mortality. Associations of NDVI with COVID-19 incidence remained in analyses stratified by urban-rural status or population density, indicating that our associations are not a result of differences in urban-rural COVID-19 incidence or testing rates. We adjusted for several potentially important confounders, such as proportion Black residents, population density, and days since first COVID-19 case. We note that NDVI was moderately positively (Spearman rho > 0.40) correlated with % less than high school education, % Black residents, % current smokers, and PM_2.5_, while these variables were all positively associated with COVID-19 incidence and mortality. Further, sensitivity analyses showed that associations were robust to exclusion of counties with 10 or fewer COVID-19 cases, excluding all counties comprising the New York metropolitan area and additional adjustment for physical distance closures and potential spatial clustering.

We acknowledge that this study has several limitations. This is an ecological study with aggregated data on county level. Ecological designs should not be used to make inferences about individual risks even though they are valid for hypothesis-generating purposes. Publicly available COVID-19 outcome data was only available at county level, while COVID-19 incidence and mortality, and sociodemographic characteristics likely vary at a smaller spatial scale. (Villeneuve and Goldberg 2020) COVID-19 events are not independent and likely cluster over time and space which may have resulted in biased effect estimates. (Villeneuve and Goldberg 2020) Although we adjusted for several important confounders, such as days since first COVID-19 case reported and days since stay-at-home order, it is possible that there is residual confounding by these factors. Days since stay-at-home order is based on the start date of the issuance of the order. However, in several states the stay-at-home order was ended/relaxed in (the end of) April or May (earlier than June 7). Further, there are other state-level physical distance closures (e.g. day cares, K-12 schools, gyms) that we did not take into account. As additional adjustment for days since non-essential business closure and days since nursing home visitor ban did not affect our associations, we do not think that adjustments for additional closures would greatly impact our findings. We also note that physical distance closures and face coverings requirements could differ between counties within a state. We used a county-level vegetation index as a proxy for green space access, which does not distinguish whether vegetation represents urban parks, forests, agricultural land, or overgrown vacant lots. Detail on park amenities, vegetation species or typology, and park usage during the COVID-19 pandemic were unavailable at the time of data collection, but would add to future analyses. Another major limitation is the underreporting of COVID-19 cases and deaths. Widespread testing has been limited in most areas of the US and differences in testing availability might differ between counties and could have changed over time due to additional resources and increased recognition of the disease.

Despite these limitations, our findings suggest that exposures to greenness had beneficial impacts on county-level incidence of COVID-19 in the US and may have reduced county-level COVID-19 mortality rates, especially in areas of higher population density. Although casual relationships cannot be drawn from ecological studies, our findings imply that keeping parks open, maintaining funding for parks in light of coming surges of COVID-19 and future pandemics may have important public health benefits.

**Table 1.** Descriptive statistics of the full cohort (n = 3,089 U.S. counties) ^a^.

| Variable | Median (IQR) |
| --- | --- |
| COVID-19 death rate (per 100,000) | 2.8 (13.4) |
| COVID-19 incidence rate (per 100,000) | 163.5 (330.2) |
| NDVI (April-May 2020) | 0.44 (0.27) |
| NDVI (June-August 2019) | 0.63 (0.21) |
| Population density (person/sq. mi.) | 60.6 (208.7) |
| % in poverty | 9.2 (6.1) |
| % owner occupied housing | 76.7 (9.3) |
| % less than high school education | 19.1 (13.2) |
| % Black | 1.4 (7.8) |
| % Hispanic | 3.1 (5.8) |
| % 65 years of age | 15.6 (5.0) |
| % 45-64 years of age | 26.5 (2.9) |
| % 15-44 years of age | 37.9 (5.4) |
| Median home value (\$1,000) | 110.3 (67.7) |
| Median household income (\$1,000) | 47.7 (15.2) |
| % Obese | 33.1 (7.2) |
| % Smoke | 17.0 (4.8) |
| Days since stay-at-home order | 68 (74) |
| Days since non-essential businesses closure | 69 (74) |
| Days since nursing homes visitor ban | 62 (83) |
| Days since first case | 74 (13) |
| Rate of hospital beds (per 100,000) | 50 (173) |
| Rate of tests (per 100,000) | 5670.7 (2394.6) |
| Average summer temperature (K) | 303.3 (5.0) |
| Average winter temperature (K) | 280.2 (10.5) |
| Average summer relative humidity (%) | 91.3 (6.7) |
| Average winter relative humidity (%) | 88 (5.6) |
| PM<sub>2.5</sub> (µg/m<sup>3</sup>) | 8.8 (4.1) |
| Urban counties [NCHS classification ≤4 (n)] | 1149 |
| Counties with issuance of stay-at-home order (n) | 2196 |
| Counties with 10< cases (n) | 2297 |

<sup>a</sup> The number of COVID-19 cases and deaths are based on data from March 22, 2020 through June 7, 2020.

## Acknowledgement

This study was funded by R01HL150119, R01ES028033 and P30 ES000002.

