## Supplemental Table 1, 2 and 3 for "County-level exposures to greenness and associations with COVID-19 incidence and mortality in the United States"

**Table S1. Data sources used in this study.**

|  | **Data source** | **Description** |
| --- | --- | --- |
| **Outcome** | Johns Hopkins University the Center for Systems Science and Engineering (JHU-CSSE) Coronavirus Resource Center (https://coronavirus.jhu.edu/) | Cumulative county-level COVID-19 cases and death (count up to and including June 7, 2020) |
| **Exposure** | Landsat 8 NDVI via Google Earth Engine (https://developers.google.com/earth-engine/datasets/catalog/LANDSAT_LO08_C01_T1?hl=en) | NDVI based on Landsat 8 images from April 1, 2020 up to May 31, 2020 (NDVI April-May) and from June 1, 2020 up to August 31, 2019 (NDVI summer) at a 30 meter spatial resolution |
| **Confounders** | US Census/American Community Survey (https://www.census.gov/progra ms-surveys/acs/data.html) | County-level socioeconomic and demographic variables for 2000‒2010 |
|  | Robert Wood Johnson Foundation County Health Rankings (https://www.countyhealthranki ngs.org/) | County-level behavioral risk factor variables for 2011 |
|  | JHU-CSSE Coronavirus Resource Center | Time since first reported COVID-19 case |
|  | Raifman et al, Boston University School of Public Health, COVID-19 United States state policy database (www.tinyurl.com/statepolicies) | Days since issuance of stay-at-home order, days since non-essential businesses closure, days since nursing home visitor ban |
|  | Homeland Infrastructure Foundation-Level Data (HIFLD) (https://hifldgeoplatform.opendata.arcgis.co m/datasets/hospitals) | County-level number of hospital beds in 2019 |
|  | Gridmet via Google Earth engine (https://developers.google.com/e arthengine/datasets/catalog/IDAHO _EPSCOR_GRIDMET) | 4 km × 4 km temperature and relative humidity predictions, summer and winter averaged across the period 2000‒2016 and averaged across grid cells in each county |
|  | Atmospheric Composition Analysis Group (https://sites.wustl.edu/acag/) | 0.01° × 0.01° grid resolution PM2.5 prediction, averaged across the period 2000‒2016 and averaged across grid cells in each county |
|  | The COVID tracking project (https://covidtracking.com/) | State-level number of COVID-19 tests performed up to and including June 7, 2020 |

**Table S2. Mortality rate ratios (MRR), Incidence Rate Ratios (IRR), 95% confidence intervals (CI) for all variables in the main analysis for COVID-19 mortality and COVID-19 cases.**

| **Variable** | **MRR (95% CI)** | **IRR (95% CI)** |
| --- | --- | --- |
| NDVI (April-May) | 0.99 (0.94, 1.05) | 0.94 (0.90, 0.97) |
| Population density (Q2) | 0.94 (0.73, 1.20) | 0.99 (0.86, 1.14) |
| Population density (Q3) | 0.88 (0.68, 1.15) | 0.86 (0.74, 1.00) |
| Population density (Q4) | 0.79 (0.60, 1.03) | 0.72 (0.62, 0.85) |
| Population density (Q5) | 0.86 (0.64, 1.17) | 0.74 (0.62, 0.90) |
| % in poverty | 1.05 (0.97, 1.14) | 1.02 (0.97, 1.07) |
| log(Median home value) | 1.17 (1.01, 1.34) | 1.08 (0.99, 1.18) |
| log(Median household income) | 1.20 (1.05, 1.37) | 1.11 (1.02, 1.21) |
| % owner occupied housing | 1.11 (1.03, 1.20) | 1.05 (0.99, 1.10) |
| % less than high school education | 1.20 (1.09, 1.32) | 1.11 (1.05, 1.18) |
| % Black | 1.51 (1.40, 1.64) | 1.36 (1.29, 1.45) |
| % Hispanic | 1.05 (0.96, 1.16) | 1.27 (1.18, 1.35) |
| % 65 years of age | 1.05 (0.93, 1.18) | 0.86 (0.80, 0.92) |
| % 15-44 years of age | 0.80 (0.68, 0.93) | 0.91 (0.85, 0.98) |
| % 45-64 years of age | 0.79 (0.70, 0.88) | 0.84 (0.79, 0.90) |
| % Obese | 0.97 (0.90, 1.04) | 0.95 (0.90, 0.99) |
| % Smoke | 1.12 (0.98, 1.27) | 1.17 (1.08, 1.27) |
| Days since stay-at-home order (state-level) | 1.18 (0.91, 1.54) | 1.03 (0.89, 1.19) |
| Days since first case | 2.42 (2.07, 2.83) | 1.86 (1.75, 1.97) |
| Rate of hospital beds | 1.00 (0.93, 1.08) | 1.04 (1.00, 1.08) |
| Average summer temperature | 1.11 (0.94, 1.31) | 1.23 (1.10, 1.37) |
| Average winter temperature | 0.89 (0.71, 1.12) | 0.67 (0.58, 0.78) |
| Average summer relative humidity (%) | 0.94 (0.79, 1.11) | 1.08 (0.97, 1.20) |
| Average winter relative humidity (%) | 0.96 (0.86, 1.07) | 0.97 (0.91, 1.05) |
| PM_2.5_ (µg/m^3^) | 1.11 (1.05, 1.17) | 1.06 (1.02, 1.10) |
| Rate of tests ^a^ | - | 1.14 (1.01, 1.27) |

*^a^ Rate of tests was included in the main analysis for COVID-19 cases but not in the main analysis for COVID-19 deaths. Results of the model including rate of tests for COVID-19 deaths can be found in figure S4.*

**Table S3. Association of April-May and Summer (June-August) NDVI with COVID-19 incidence and COVID-19 mortality in the full cohort, in urban counties and in rural counties ^a^.**

| **Outcome** | **NDVI** | **Full cohort** | **Urban counties** | **Rural counties** |
| --- | --- | --- | --- | --- |
|  |  | **HR (95% CI)** | **HR (95% CI)** | **HR (95% CI)** |
| **Incidence** | **April-May** | 0.94 (0.90, 0.97) | 0.92 (0.88, 0.96) | 0.97 (0.92, 1.02) |
|  | **Summer** | 0.98 (0.95, 1.02) | 0.96 (0.91, 1.01) | 1.02 (0.97, 1.07) |
| **Mortality** | **April-May** | 0.99 (0.94, 1.05) | 0.95 (0.89, 1.01) | 0.99 (0.91, 1.08) |
|  | **Summer** | 0.97 (0.92, 1.02) | 0.94 (0.88, 1.00) | 0.96 (0.88, 1.04) |

^a^ Associations are expressed per 0.1 unit increase in NDVI.
