## Supplemental figures for "County-level exposures to greenness and associations with COVID-19 incidence and mortality in the United States"

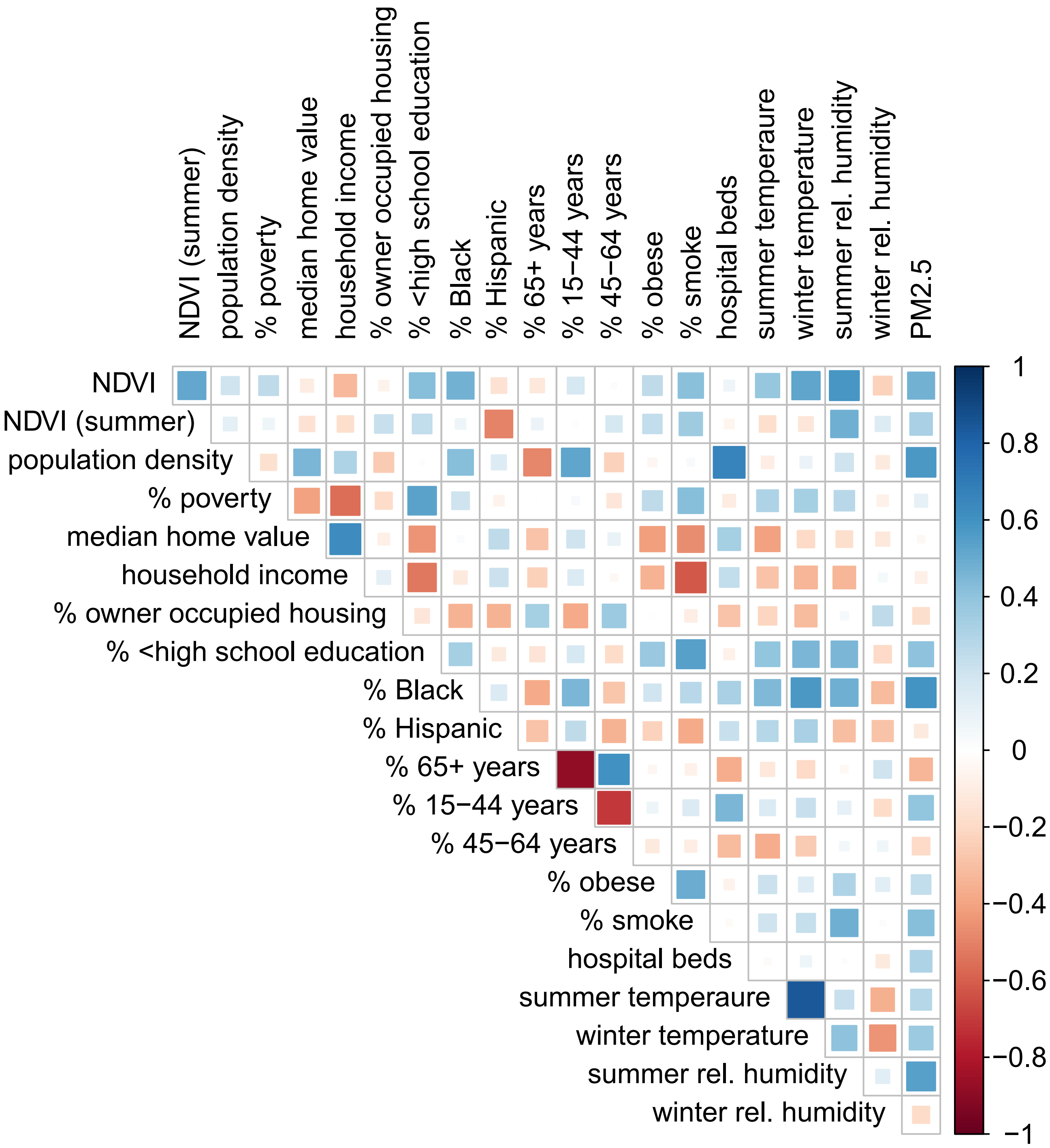


Figure S1. Spearman correlation between NDVI, US census variables, BRFSS variables (% obese, % smoke), # hospital beds per county, temperature, relative humidity and PM2.5 [a].

[a] Each cell is colored blue or red depending on the sign of the correlation and with the intensity of color scaled.

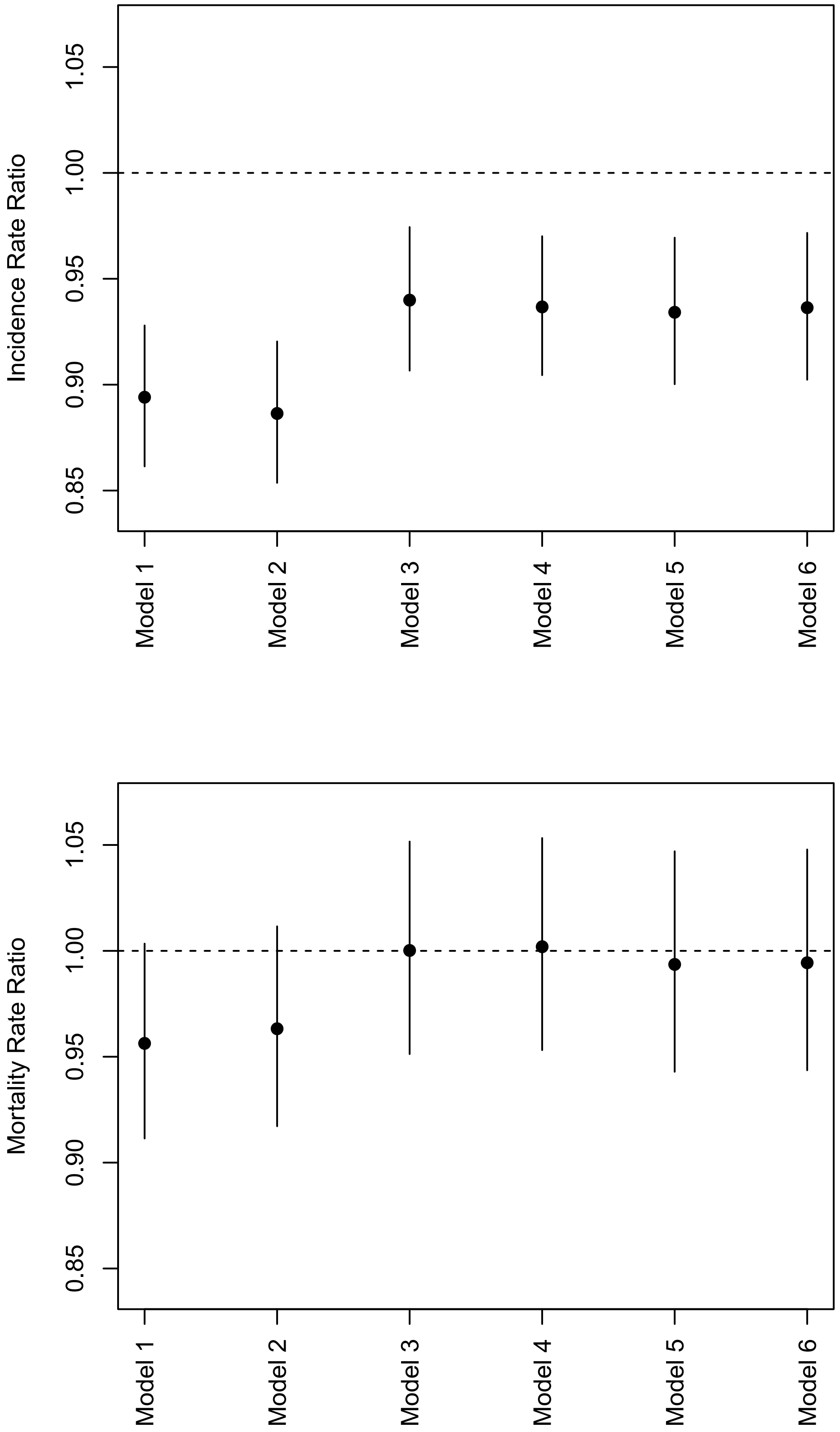


Figure S2. Associations of NDVI with COVID-19 incidence and COVID-19 mortality in models with increasing adjustment for covariates. Model 1: NDVI + population size offset + random intercept by state, Model 2: Model 1 + population density, Model 3: Model 2 + county-level SES covariates and BRFSS covariates, Model 4: Model 3 + date since first COVID-19 case + date since issuance of stay-at-home order + number of hospital beds per unit population, Model 5: Model 4 + temperature + relative humidity + PM2.5, Model 6: Model 5 + number of test per unit population [a].

[a] Associations are expressed per 0.1 unit increase in NDVI.

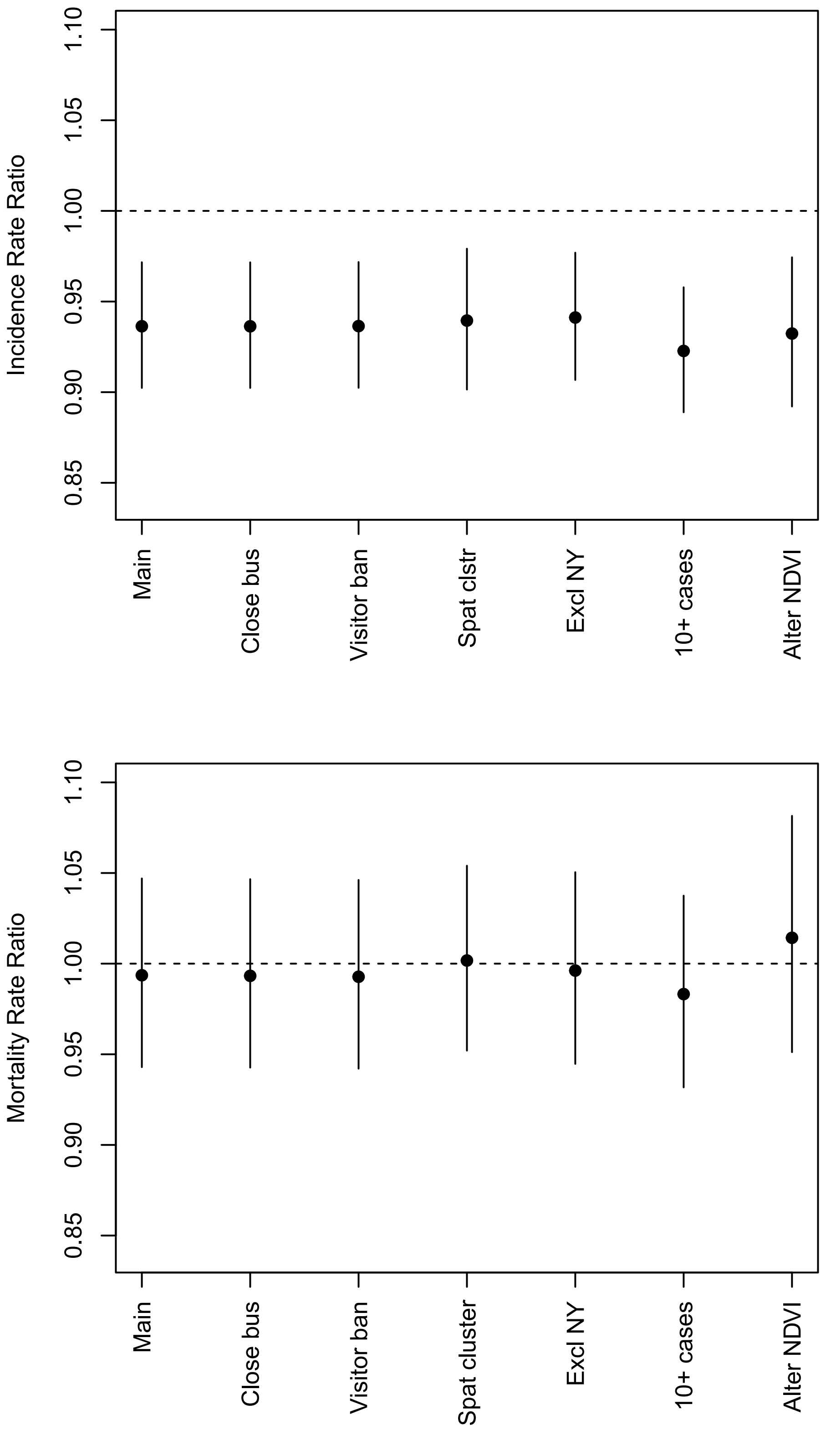


Figure S3. Associations of NDVI with COVID-19 incidence and COVID-19 mortality in sensitivity analyses. Main = main model, Closed bus = additional adjustment for days since closure of non-essential businesses, Visitor ban = additional adjustment for days since nursing home visitor ban, Spat cluster = additional adjustment for longitude and latitude of the centroid of each county, Excl NY = all counties comprising the New York metropolitan area were excluded from the dataset, Excl 10 cases = all counties with less than 10 cases were excluded from the dataset, Alter NDVI = alternative procedure for calculating NDVI (excluding negative values) [a].

[a] Associations are expressed per 0.1 unit increase in NDVI.
